# Association between daily sitting time and urinary leakage: double mediation effects of inflammation and physical activity

**DOI:** 10.1101/2025.04.22.25326195

**Authors:** Chunling Huang, Li Wang, Huiwen Hu, Bin Yu

**Author notes:** Corresponding author: Bin Yu,; Huiwen Hu. Chunling Huang and Li Wang contribute equally to this paper.

## Abstract

**Highlights:** Longer daily sitting time is associated with increased risk of urinary leakage Physical activity mitigates the urinary leakage risk from prolonged sitting Inflammation mediates the link between sitting and urinary leakage

**Background:** Sedentary behavior (SB), recognized as a health risk, may contribute to urinary leakage (UL). This study examined the association between daily sitting time (DST) and UL risk, investigating inflammatory markers as mediators and leisure-time physical activity (LTPA) as a moderator.

**Methods:** We conducted a cross-sectional analysis of data from the National Health and Nutrition Examination Survey (NHANES) spanning 2007 to 2020, involving 23,417 participants. Information on UL, DST, Dietary Inflammatory Index (DII), Body Mass Index (BMI), and Neutrophil-to-Lymphocyte Ratio (NLR) was collected through standardized questionnaires and 24-hour recall interviews. Logistic regression models were employed to examine the relationship between DST, LTPA, and UL, adjusting for relevant covariates. Mediation analysis was conducted to assess the potential mediating effects of DII, BMI, and NLR on the association between DST and UL.

**Results:** The overall prevalence of UL among participants was 29.19%. Our findings revealed a significant positive association between longer DST and UL risk. Notably, individuals engaging in adequate LTPA demonstrated an attenuated association between DST and UL. Furthermore, mediation analysis identified significant parallel double mediating effects of DII-BMI and BMI-NLR on the relationship between DST and UL. However, no significant mediating effect was observed for DII-NLR, this suggests that DII, BMI, and NLR might jointly mediate the impact of DST on UL risk.

**Conclusions:** These findings suggest that prolonged sitting may elevate UL risk through inflammatory mechanisms, and that LTPA may mitigate this risk. Further research is needed to validate these results and explore the underlying biological pathways.

## Introduction

Urinary leakage (UL), also known as urinary incontinence (UI), is a widespread health concern characterized by the involuntary loss of urine. In the United States, this condition affects a substantial portion of the population.^1^ According to estimates, the prevalence ranges from 9.3% to 30.8% among women and 2.6% to 20.9% among men, with a notable increase in prevalence observed with advancing age.^2^ UL can have a profound impact on an individual’s life, leading to compromised psychosocial well-being, diminished self-confidence, and reduced social interactions and interpersonal relationships.^3^ Therefore, it is essential to identify the risk factors and mechanisms underlying UL in order to provide comprehensive disease management.

Sedentary behavior (SB) are recognized as independent risk factors for numerous health-related issues. SB refers to any waking activity performed while sitting, reclining, or lying down that expends no more than 1.5 metabolic equivalents (METs).^4^ SB has been identified as a risk factor for various conditions, including diabetes, obesity, cardiovascular disease, cancer, and dementia. A cross-sectional study involving 150 women at a urology center revealed a link between SB and the occurrence of UI in postmenopausal women.^5,6^ Additionally, an analysis by Roig et al. of women aged 60 years and older from the NHANES database demonstrated a significant association between increased sedentary time and UI in older women, suggesting that reducing prolonged sitting time may be an effective intervention for lowering UI risk.^7^ These studies indicate a potential association between SB and UI. However, the specific reasons and key factors behind prolonged sitting leading to UL are unclear, and elucidating its specific mechanism is particularly important.

Sedentary behavior is strongly associated with the development of systemic low-grade inflammation,^8,9^ potentially mediated by metabolic dysregulation such as hypertriglyceridemia, elevated low-density lipoprotein cholesterol, insulin resistance, and poor glycemic control.^10,11^ Studies have shown that chronic, low-grade inflammation is prevalent in individuals who maintain sedentary lifestyles. According to research, inflammation can change with changes in dietary components, and DII is related to systemic inflammatory reactions.^12^ In American women under the age of 65, DII is positively correlated with UI.^13^ Furthermore, accumulating evidence suggests a significant positive correlation between a pro-inflammatory systemic environment and the risk of developing UI, indicating that elevated inflammatory states may increase the likelihood of UI.^14^ In recent years, leisure-time physical activity (LTPA) has become a hot topic, with research showing that LTPA can improve chronic inflammatory status within the body. For example, consistent and structured exercise training can reduce the production of inflammatory cytokines and downregulate the cell-surface expression of TLR4 on monocytes.^15^ However, there is a lack of systematic research on the relationship between prolonged sitting, chronic inflammation, and LTPA.

Based on this background, we hypothesize that inflammation may serve as a crucial link between SB and UL. The NHANES is an ongoing, nationally representative survey designed to assess the health and nutritional status of the US population. This survey encompasses a broad range of health-related aspects, including demographics, socioeconomic factors, dietary and health-related behaviors, and utilizes interviews, physical examinations, and laboratory tests to collect data.^16^ This study aims to utilize the NHANES database to investigate the relationship between DST and UL in US adults, with a particular focus on exploring the mediating role of inflammation. We believe that the findings of this study will provide valuable insights and guidance for the prevention of UL.

## Materials and Methods

### Study design and participants

The NHANES database is a population-based cross-sectional survey program conducted by the Centers for Disease Control and Prevention in the United States. It aims to assess the health and nutritional status of the American population, estimate the distribution of certain diseases and risk factors within the US population and specific subgroups, monitor trends in risk behaviors and environmental exposures, and explore emerging public health issues and new technologies. This study utilized data from the NHANES database from 2007 to 2020. Initially, 66,148 people were included, after excluding demographic data with missing values (n=42,731), questionnaire data with missing values (n=1,921), and variable data with missing values (n=1,328), our final analysis included 20,168 participants who met the conditions, as shown in Figure 1. All NHANES protocols received approval from the Ethics Review Board of the National Center for Health Statistics, and written informed consent was obtained from each participant.

**Fig 1.**
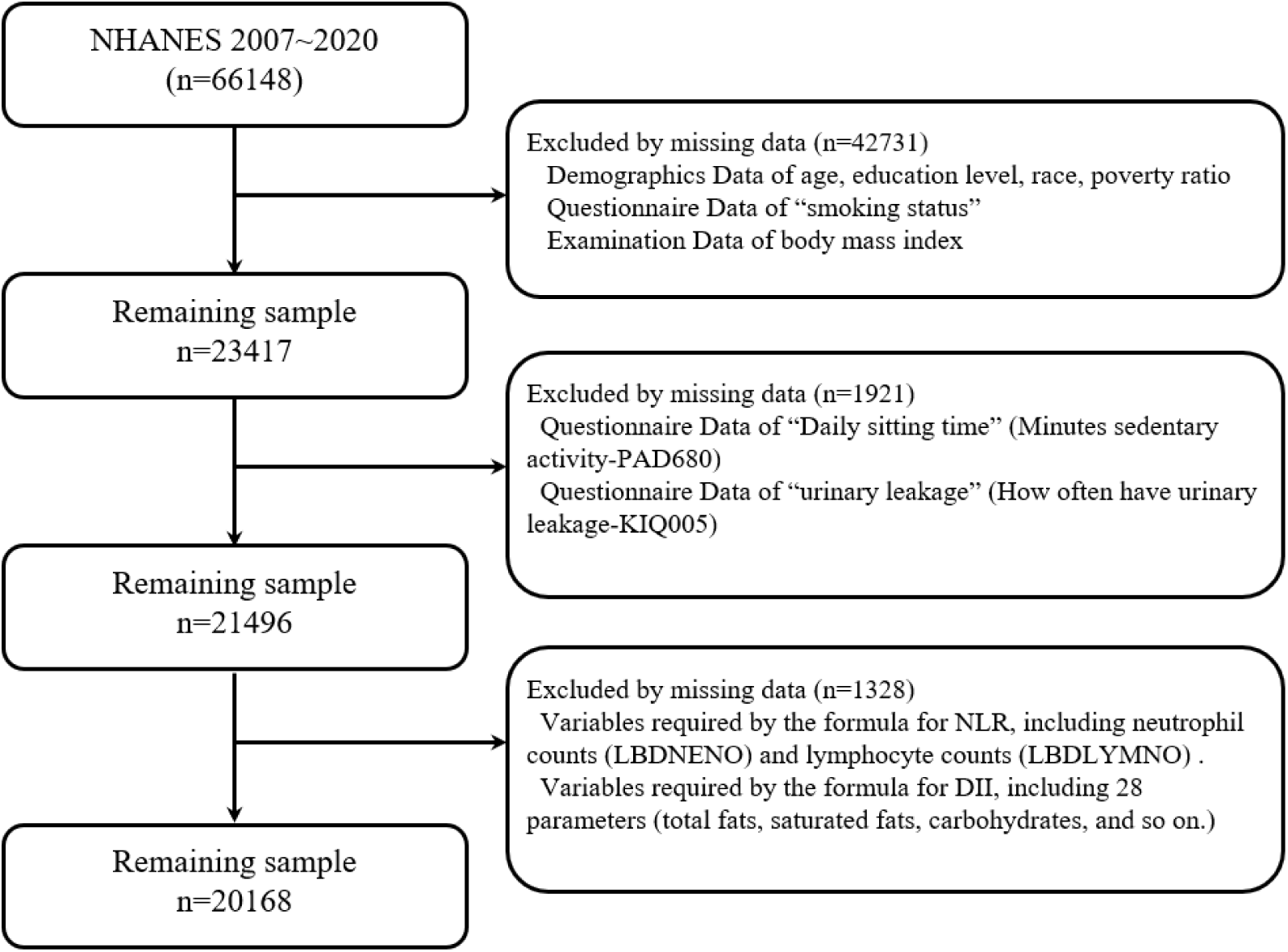
Flowchart of participant selection in NHANES 2007–2020.

### Main variables

#### 1. Daily sitting time (DST)

The term “daily sitting time (DST)” was defined in the Questionnaire Data of Physical Activity (PAQ) survey as “Daily sitting time (Question number: PAD680): How much time do you usually spend sitting on a typical day?” The question is about sitting at work, at home, getting to and from places, or with friends, including time spent sitting at a desk, traveling in a car or bus, reading, playing cards, watching television, or using a computer. Do not include time spent sleeping. For further analysis, the time was converted into hours. According to the method by Li et al., DST was categorized into four groups: G1 (less than 4 hours per day), G2 (4 to 6 hours per day), G3 (6 to 8 hours per day), and G4 (8 hours or more per day). In the subsequent analysis, a DST of less than 4 hours per day was used as the reference. ^16^

#### 2. UL

The definition of UL came from the Questionnaire Data in the Kidney Conditions - Urology (KIQ) survey. When asked “How often have urinary leakage? (Question number: KIQ005)”, women who answer “never” are considered not to have UL. Other responses are classified as having urinary leakage and are grouped based on their answers (UL1: Less than once a month. UL2: A few times a month. UL3: A few times a week. UL4: Every day and/or night).

#### 3. NLR

NLR is a blood indicator used to assess the body’s inflammatory status and immune response. It can be obtained from the Laboratory Data in the Complete Blood Count (CBC). The formula for calculation is: NLR = neutrophil (LBDNENO) / lymphocyte (LBDLYMNO).

#### 4. DII

Extracted data from the NHANES database using the 24-hour dietary recall questionnaire included 28 parameters to calculate the DII, such as total fat, saturated fat, carbohydrates, protein, alcohol, monounsaturated fat, fiber, cholesterol, n-3 fatty acids, polyunsaturated fat, vitamin B12, vitamin E, n-6 fatty acids, niacin, vitamin A, thiamine (vitamin B1), beta-carotene, folic acid, riboflavin (vitamin B2), vitamin B6, vitamin C, vitamin D, selenium, iron, magnesium, zinc, caffeine, and energy.^17^

#### 5. LTPA

Extract the Physical Activity (PAQ) data from the NHANES database Questionnaire Data. Collect PAD660 - Minutes of vigorous recreational activities and PAD675 - Minutes of moderate recreational activities. Calculate LTPA using the formula from the literature: the total time spent on LTPA was calculated as minutes of moderate-intensity recreational activities plus twice the minutes of vigorous-intensity recreational activities. According to the 2018 Physical Activity Guidelines for Americans: individuals with no LTPA, those with LTPA greater than 0 but less than 150 minutes/week, and those with LTPA greater than or equal to 150 minutes/week are classified as inactive, insufficiently active, and sufficiently active, respectively.^18^

### Others Covariate

BMI is defined as: Examination data includes BMI - Body Mass Index (kg/m²). Classification criteria: (under/normal weight: <25 kg/m², overweight: 25 to 30 kg/m², obesity: >30 kg/m²). Additionally, some covariates from the NHANES database were used, including age, race, education level, poverty-income ratio, and smoking.

### Statistical Analysis

All statistical analyses were conducted following the guidelines provided by the Centers for Disease Control and Prevention (CDC), and appropriate sampling weights were applied for participant recruitment. Baseline characteristics of the included population were described and grouped by different DST. Continuous variables were summarized using mean (SD), while categorical variables were presented as percentages.

To analyze the association between DST and UL, we employed a crude model and adjusted models in logistic regression analysis: Model 1 without adjustments; Model 2 was additionally adjusted for age, gender, race, education level, poverty, and smoking status; Model 3 was further adjusted for BMI, NLR, and DII. The strength of associations was estimated using odds ratios (ORs) and their corresponding 95% confidence intervals (CIs).

Mediation analysis was performed using the mediation package in R software. Through mediation analysis, we can calculate how much of an effect needs to be mediated. This is an ideal strategy to elucidate pathways and provide statistical evidence for mechanistic analysis. In this study, the direct effect represented the correlation between DST, LTPA, and UL; the indirect effect, which was mediated by inflammatory indicators, represented the correlation between DST, LTPA, and UL; the mediation proportion represented the percentage of mediation effect. All analyses were performed using the R software (version 4.2.0, http://www.R-project.org, The R Foundation). *P* < 0.05 indicated statistically significant differences.

## Results

### Population characteristics

From 2007 to 2020, a total of 66,148 individuals were initially recruited for the survey. After excluding those with missing data on age, education level, DST, UL and other factors, 20,168 participants were included in our study (Figure 1). Statistical analysis of these baseline characteristics revealed that increased DST was significantly associated with higher levels of inflammation markers, compared to group G1, groups G2, G3, and G4 showed significantly higher Systemic Inflammation Index (SII), Neutrophil-to-Lymphocyte Ratio (NLR), as well as elevated white blood cell (WBC) count and neutrophils (Ne) values, indicating a state of chronic inflammation. Besides, the overall incidence of UL was 29.19%. Compared to the group with G1 (26.02%), groups G2, G3, and G4 showed higher incidence and severity of UL. As DST increased, the DII gradually decreased, while the proportion below zero significantly rose (all *P*<0.01). Detailed data can be found in Table 1.

**Table 1.** Characteristics of Participants Classified by DST: NHANES 2007-2020.

| Variable Names | Overall | Daily sitting time (h) |  |  |  | p-value |
| --- | --- | --- | --- | --- | --- | --- |
|  |  | <4 | 4~6 | 6~8 | >=8 |  |
| **n** | 20168 | 5449 | 4783 | 3204 | 6732 |  |
| **Age (year)** | 49.25 (17.66) | 47.88 (16.57) | 49.8 (18.03) | 50.73 (18.72) | 49.26 (17.66) | <0.01 |
| **gender (%)** |  |  |  |  |  |  |
| Female | 10106 (50.11) | 2761 (50.67) | 2344 (49.01) | 1580 (49.31) | 3421 (50.82) | 0.160 |
| Male | 10062 (49.89) | 2688 (49.33) | 2439 (50.99) | 1624 (50.69) | 3311 (49.18) |  |
| **Race (%)** |  |  |  |  |  |  |
| Mexican American | 3030 (15.02) | 1353 (24.83) | 712 (14.89) | 359 (11.20) | 606 (9.00) | <0.001 |
| Other Hispanic | 2059 (10.21) | 758 (13.91) | 503 (10.52) | 282 (8.80) | 516 (7.66) |  |
| Non-Hispanic White | 9159 (45.41) | 1984 (36.41) | 2217 (46.35) | 1591 (49.66) | 3367 (50.01) |  |
| Non-Hispanic Black | 3996 (19.81) | 971 (17.82) | 944 (19.74) | 627 (19.57) | 1454 (21.60) |  |
| Other Race | 1924 (9.54) | 383 (7.03) | 407 (8.51) | 345 (10.77) | 789 (11.72) |  |
| **Poverty income ratio** | 2.52 (1.63) | 2.13 (1.5) | 2.44 (1.58) | 2.55 (1.64) | 2.87 (1.68) | <0.01 |
| **Education level (%)** |  |  |  |  |  |  |
| Less than 9th grade | 1933 (9.58) | 895 (16.43) | 449 (9.39) | 232 (7.24) | 357 (5.30) | <0.001 |
| 9-11th grade | 2829 (14.03) | 991 (18.19) | 739 (15.45) | 408 (12.73) | 691 (10.26) |  |
| High school graduate | 4585 (22.73) | 1351 (24.79) | 1165 (24.36) | 750 (23.41) | 1319 (19.59) |  |
| Some college | 6018 (29.84) | 1440 (26.43) | 1443 (30.17) | 1020 (31.84) | 2115 (31.42) |  |
| College graduate or above | 4803 (23.81) | 772 (14.17) | 987 (20.64) | 794 (24.78) | 2250 (33.42) |  |
| **BMI (kg/m<sup>2</sup>)** |  |  |  |  |  |  |
| Normal weight (%) | 5764 (28.58) | 1634 (29.99) | 1383 (28.91) | 926 (28.90) | 1821 (27.05) | <0.001 |
| Overweight (%) | 6718 (33.31) | 1941 (35.62) | 1672 (34.96) | 1002 (31.27) | 2103 (31.24) |  |
| Obesity (%) | 7686 (38.11) | 1874 (34.39) | 1728 (36.13) | 1276 (39.83) | 2808 (41.71) |  |
| **UL(%)** |  |  |  |  |  |  |
| No | 14281 (70.81) | 4031 (73.98) | 3435 (71.82) | 2197 (68.57) | 4618 (68.60) | <0.001 |
| Yes | 5887 (29.19) | 1418 (26.02) | 1348 (28.18) | 1007 (31.43) | 2114 (31.40) |  |
| **KIQ005 (%)** |  |  |  |  |  |  |
| A few times a month | 1867 (9.26) | 484 (8.88) | 449 (9.39) | 303 (9.46) | 631 (9.37) | <0.001 |
| A few times a week | 980 (4.86) | 246 (4.51) | 193 (4.04) | 175 (5.46) | 366 (5.44) |  |
| Every day and/or night | 1093 (5.42) | 219 (4.02) | 239 (5.00) | 210 (6.55) | 425 (6.31) |  |
| Less than once a month | 1947 (9.65) | 469 (8.61) | 467 (9.76) | 319 (9.96) | 692 (10.28) |  |
| Never | 14281 (70.81) | 4031 (73.98) | 3435 (71.82) | 2197 (68.57) | 4618 (68.60) |  |
| **DII(%)** |  |  |  |  |  |  |
| <0 | 4477 (22.20) | 1158 (21.25) | 1041 (21.76) | 694 (21.66) | 1584 (23.53) | 0.013 |
| ≥0 | 15691 (77.80) | 4291 (78.75) | 3742 (78.24) | 2510 (78.34) | 5148 (76.47) |  |
| **LTPA(%)** |  |  |  |  |  |  |
| Insufficiently active | 985 (33.53) | 226 (29.66) | 192 (28.44) | 160 (34.78) | 407 (39.10) | <0.001 |
| Sufficiently active | 1953 (66.47) | 536 (70.34) | 483 (71.56) | 300 (65.22) | 634 (60.90) |  |
| **WBC (1000 cells/uL)** | 7.24 (2.57) | 7.14 (2.06) | 7.26 (3.26) | 7.23 (2.5) | 7.32 (2.4) | <0.01 |
| **Neutrophils (1000 cells/uL)** | 4.27 (1.83) | 4.19 (1.61) | 4.26 (2.04) | 4.29 (1.95) | 4.35 (1.78) | <0.01 |
| **Lymphocytes (1000 cells/uL)** | 2.16 (1.35) | 2.17 (0.84) | 2.19 (2.08) | 2.13 (1.05) | 2.16 (1.13) | 0.29 |
| **NLR** | 2.18 (1.22) | 2.1 (1.03) | 2.16 (1.21) | 2.23 (1.27) | 2.24 (1.33) | <0.01 |
| **SII** | 531.68 (384.72) | 516.96 (301.36) | 531.31 (518.84) | 538.12<br>(352.01) | 540.79<br>(345.62) | <0.01 |

### The relationship between DST and UL

According to the data presented in Table 2, all three models indicated a statistically significant positive correlation between DST and the risk of UL. The table listed the *P*-values, odds ratios (OR), and 95% confidence intervals (CI) for the three multivariable logistic regression models. In the unadjusted model (Model 1), compared to group G1, group G2 had a 12.0% increased risk of UL (OR=1.12, 95% CI: 1.02-1.22, *P*=0.014), group G3 had a 30% increased risk (OR=1.30, 95% CI: 1.18-1.43, *P*<0.001), and group G4 also face a 30% increased risk (OR=1.30, 95% CI: 1.20-1.41, *P*<0.001). Notably, even after adjusting for confounding factors such as age, gender, race, education, poverty, and smoking (Model 2), as well as further adjusting for BMI, NLR, and DII (Model 3), sedentary time still significantly increased the risk of UL. Trend tests indicated a significant upward trend in the risk of UL with increasing sedentary time (*P*<0.001).

**Table 2.** Multivariable-adjust ORs and 95%CI of DST and UL.

| Daily sitting time | <sup>a</sup>Model 1 |  | <sup>b</sup>Model 2 |  | <sup>c</sup>Model 3 |  |
| --- | --- | --- | --- | --- | --- | --- |
|  | OR (95%CI) | p-value | OR (95%CI) | p-value | OR (95%CI) | p-value |
| **Sitting time** | 1.03 (1.02-1.04) | <0.001 | 1.04 (1.03-1.05) | <0.001 | 1.03 (1.02-1.04) | <0.001 |
| <4h | Reference |  | Reference |  | Reference |  |
| 4~6h | 1.12 (1.02-1.22) | 0.014 | 1.06 (0.97-1.17) | 0.215 | 1.03 (0.94-1.14) | 0.494 |
| 6~8h | 1.30 (1.18-1.43) | <0.001 | 1.23 (1.10-1.37) | <0.001 | 1.17 (1.05-1.30) | 0.005 |
| >=8h | 1.30 (1.20-1.41) | <0.001 | 1.30 (1.18-1.42) | <0.001 | 1.19 (1.08-1.30) | <0.001 |
| p of trend | <0.001 |  | <0.001 |  | <0.001 |  |

<sup>a</sup>Model 1 without adjustments.
<sup>b</sup>Model 2 was additionally adjusted for age, gender, race, education level, poverty, smoking status.
<sup>c</sup>Model 3 was additionally adjusted for BMI, NLR, DII.

### The double mediation effects of DII-BMI-NLR

To evaluate the association of inflammation between DST and UL risk, restricted cubic spline (RCS) analysis and mediation analysis were performed. After adjusting for age, gender, race, education, poverty, and smoking statue, there was a nonlinear positive correlation between NLR and the risk of UL caused by DST (*P*=0) by RCS analysis. As NLR increased, the risk of UL due to DST also increased (Figure 2A). Furthermore, parallel mediation analysis revealed NLR had significant mediated effects on the association of DST with UL risk, and the proportion of mediation was 3.92% (*P*=0.002) (Supplementary Figure 1). Similarly, there was a non-linear positive correlation between DII and the risk of UL caused by DST (*P*=0.04). As DII increases, it further increased the risk of UL due to DST (Figure 2B). However, RCS analysis indicated that BMI showed a linear positive correlation with the risk of UL due to DST, but no nonlinear relationship was found (Supplementary Figure 2). Subsequently, we conducted a dual mediation effect analysis, significant parallel dual mediation effects were observed for DII-BMI (Summary indirect effect=0.0042, *P*=0.0000, Figure 3A) and BMI-NLR (Summary indirect effect=0.0056, *P*=0.0000, Figure 3B), whereas DII-NLR (Summary indirect effect=0.0001, *P*=0.6475, Figure 3C) did not show such effects. It is speculated that DII-BMI-NLR might jointly mediate the risk of UL due to prolonged sitting.

**Fig. 2.**
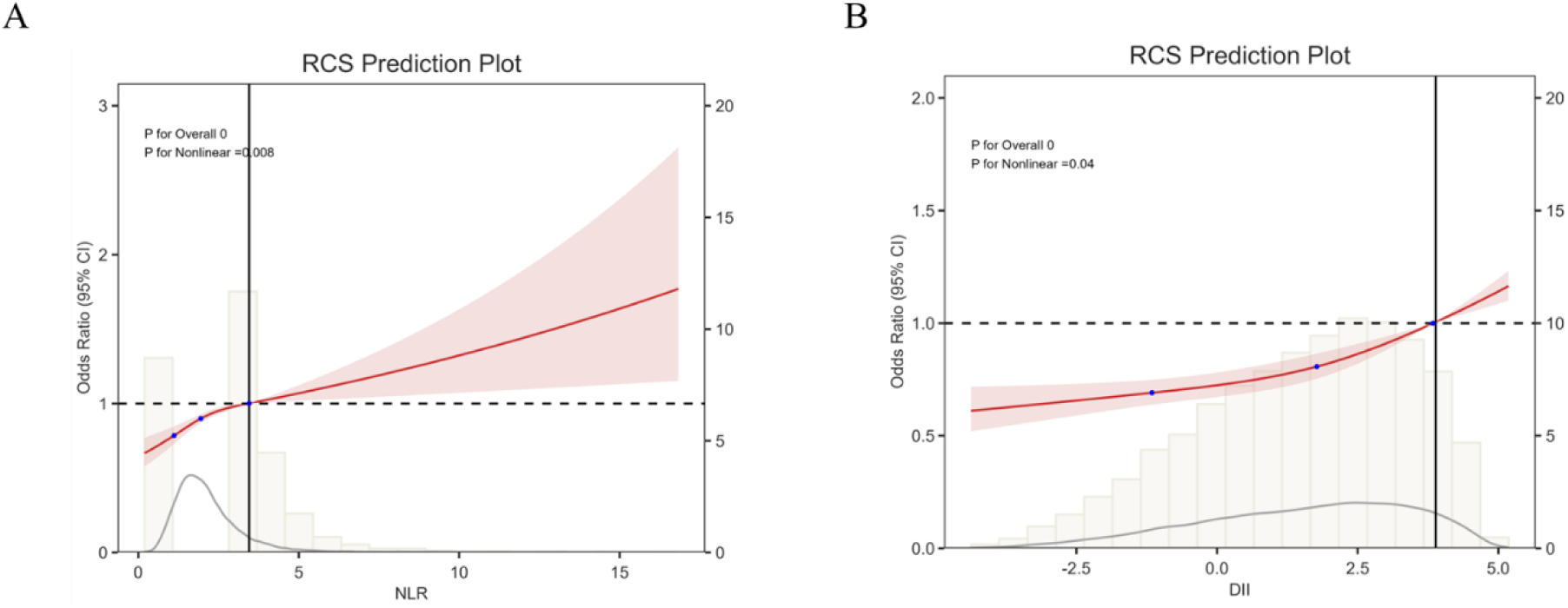
RCS revealed the association between NLR, DII, and the risk of UL caused by sedentary behavior.

**Fig. 3.**
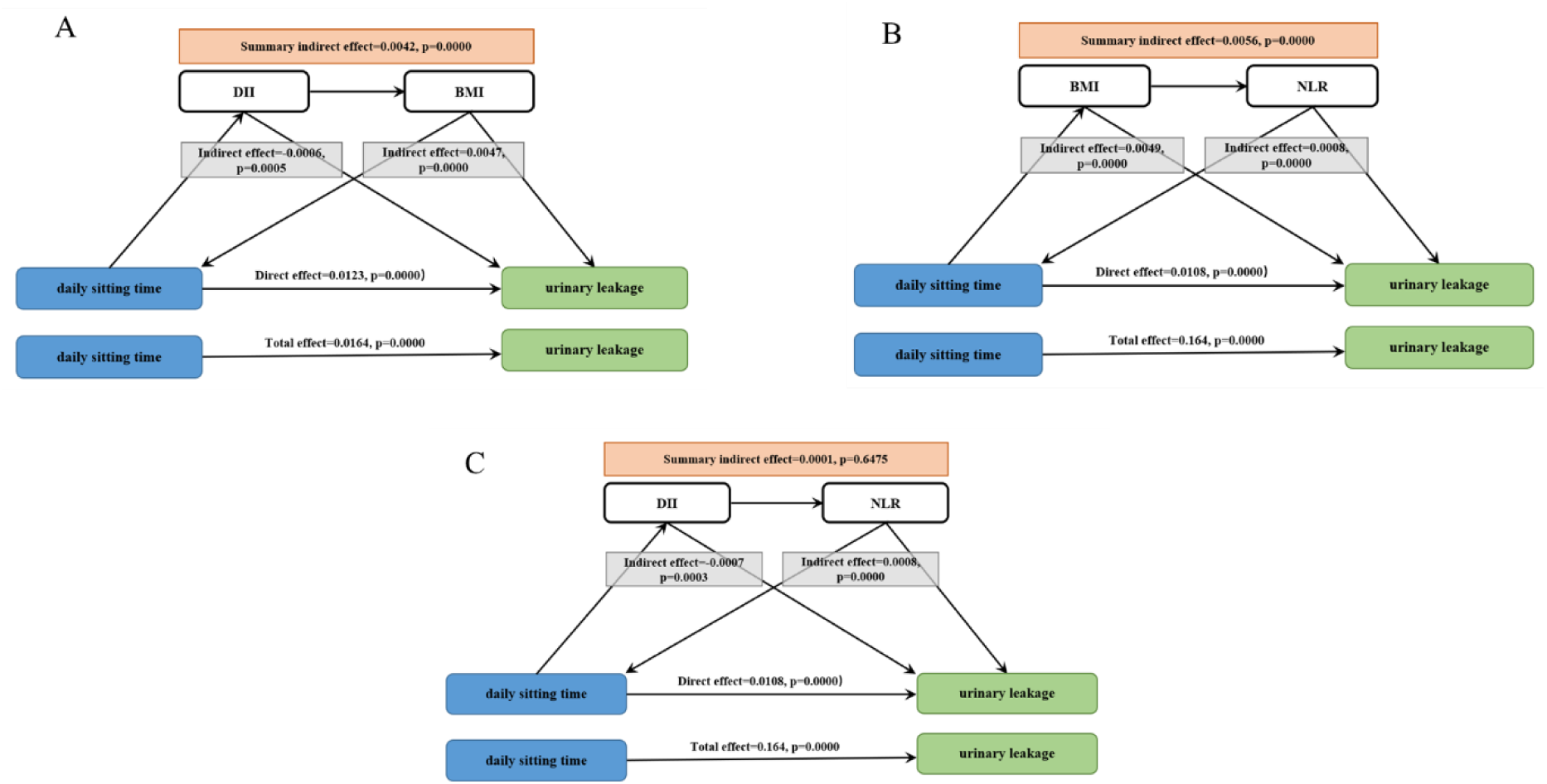
Path diagram of the double mediation analysis of DII, BMI and NLR on the relationship between DST and UL. The graphs in (A-C) represented the mediating role of DII-BMI, BMI-NLR and DII-NLR, respectively.

### LTPA improved the risk of UL caused by prolonged sitting through BMI

Current studies have shown the effectiveness of exercise interventions on UI and pelvic organ prolapse in pregnant and postpartum women.^19^ In our study, LTPA was also found to have had a linear negative correlation with the risk of UL caused by prolonged DST, but no non-linear relationship existed (Figure 4A). A double mediation analysis revealed that the LTPA-BMI had a significant parallel double mediation effect, suggesting that moderate LTPA might have reduced the risk of UL caused by prolonged DST by improving BMI (Figure 4B).

**Fig. 4.**
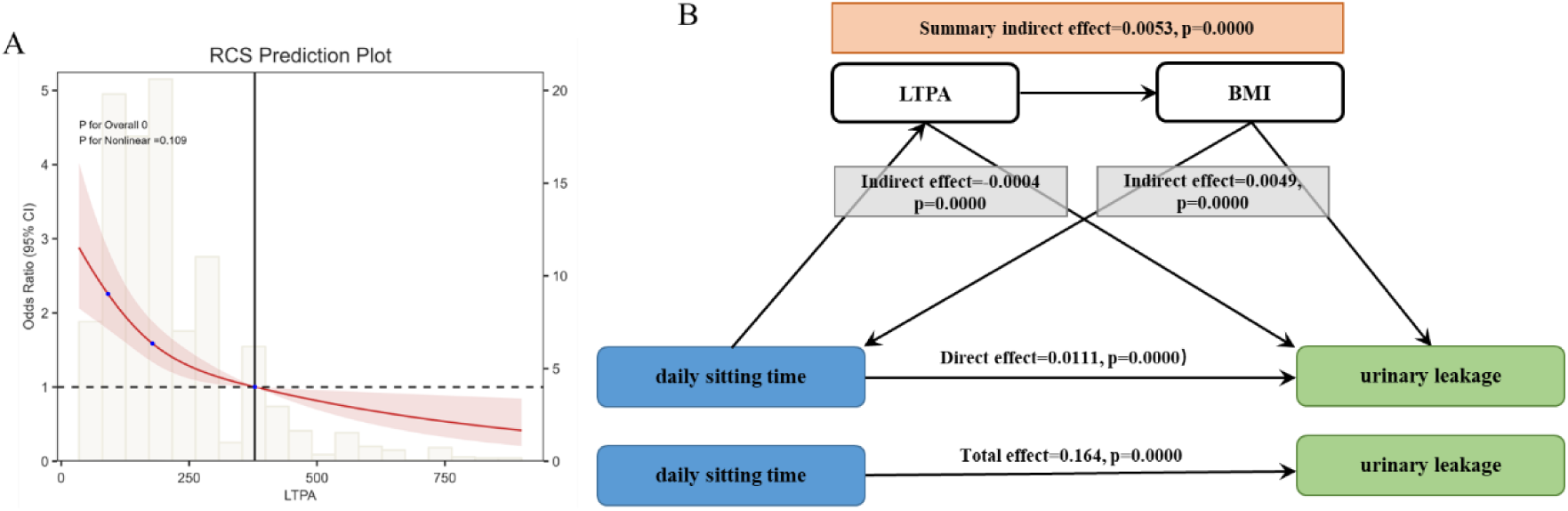
The association between LTPA and UL. (A) RCS revealed the association between LTPA and the risk of UL caused by sedentary behavior. (B) The mediating role of LTPA-BMI.

## Discussion

This research utilized data from the NHANES to investigate the association between DST and UL, and revealed meaningful and practical results. First, we observed a significant positive correlation between DST and UL, and found that inflammatory markers played a certain role in this association through their double mediation effect. Specifically, our study revealed the mediating effects of DII-BMI and BMI-NLR in the relationship between DST and UL for the first time, providing a deeper perspective on the complex mechanisms underlying this association.

Consistent with previous studies, our research demonstrated a significant association between DST and increased risk of UL, even after adjusting for confounding factors such as age, sex, race, education level, poverty, and smoking. For instance, a systematic review conducted by Faleiro et al. showed that a sedentary lifestyle and less than 150 minutes of physical activity per week are at risk of developing UI. Walking (at least 30 minutes) and physical activities (600-1,500 and 600 METs per minute per week) can prevent UI.^20^ Furthermore, prolonged sitting can lead to the accumulation of abdominal fat, increasing intra-abdominal pressure, and subsequently increasing the pressure on the bladder and pelvic muscles, further increasing the risk of UI.^21^

Further analysis revealed that inflammatory factors play a mediating role in the relationship between DST and UL risk. NLR and DII as inflammatory markers showed a non-linear positive correlation with DST, even after adjusting for confounding factors. This suggests that inflammatory reactions may be an important bridge connecting DST and UL risk. The possible mechanism is that prolonged sitting leads to chronic low-grade inflammation, characterized by elevated levels of inflammatory factors such as C-reactive protein (CRP) and tumor necrosis factor-alpha (TNF-α).^22^ These inflammatory factors can affect the pelvic muscle tissue, leading to muscle damage and subsequently increasing the risk of UL.^23^

Additionally, this study found that BMI were linearly positively correlated with the risk of UL caused by prolonged sitting, and observed a parallel double mediation effect of DII-BMI and BMI-NLR. LTPA can reduce the risk of UL caused by prolonged sitting. DII reflects the long-term impact of dietary intake on the body’s inflammatory state, with high DII diets, such as high-sugar and high-saturated fat diets, promoting the production of more inflammatory factors and exacerbating inflammatory reactions.^24^

BMI reflects the overall fat level of the body, particularly abdominal fat, which is closely related to chronic low-grade inflammation and increases intra-abdominal pressure, putting more pressure on the pelvic muscles.^25^ This suggests that improving dietary structure, controlling weight, and engaging in appropriate physical activity may be effective strategies to reduce the risk of UL in individuals with prolonged sitting.

However, there also are some limitations. First, it is a cross-sectional study, which cannot determine causality. Second, this study only included a limited number of inflammatory markers, and future studies should comprehensively evaluate the role of inflammation in the relationship between DST and UL risk. For example, considering more inflammatory factors, such as CRP and TNF-α, to comprehensively assess the inflammatory state. Additionally, the study sample was sourced from the NHANES database, which may have selection bias and information bias. Therefore, more diverse data sources are needed to support our conclusions. In summary, the study findings demonstrate a significant association between prolonged sitting time and increased UL risk, and inflammatory reactions may be a key mechanism. Future prospective studies are needed to verify these findings and explore potential intervention measures, such as randomized controlled trials, to evaluate the effectiveness of reducing sitting time, improving dietary structure, and controlling weight in reducing UL risk.

### Conclusion

Our research has revealed a strong link between prolonged sitting and an increased risk of UL. This association appears to be significantly influenced by inflammatory biomarkers, with the “dual mediation effect” highlighting the regulatory role of inflammation in this process. Encouragingly, our findings suggest that engaging in regular LTPA can help mitigate the risk of UL associated with prolonged sitting. Therefore, adopting a holistic approach that includes a balanced diet, weight management, and adequate physical activity may be key to reducing the risk of UL. These findings offer valuable insights for preventing UL and provide a foundation for future research to confirm these findings and investigate potential interventions.

## Declarations

### Ethics Approval and Consent to participate

This research was conducted independently, without patient involvement. Patients were not engaged in any stage of the research process, including the design of the study, the identification of relevant outcomes, or the interpretation of findings.

### Consent for publication

Not applicable.

### Availability of data and materials

All data supporting the findings of this study are available within the article. For any further inquiries, please contact the corresponding author.

### Authors’ contributions

CLH and BY designed the study. CLH, LW and HWH collected and analyzed data, and produced the initial draft of the manuscript. CLH, LW, HWH and BY contributed to drafting the manuscript. All authors reviewed the manuscript.

## Data Availability

All relevant data are within the manuscript and its Supporting Information files

---

**Supplementary Fig.1.**
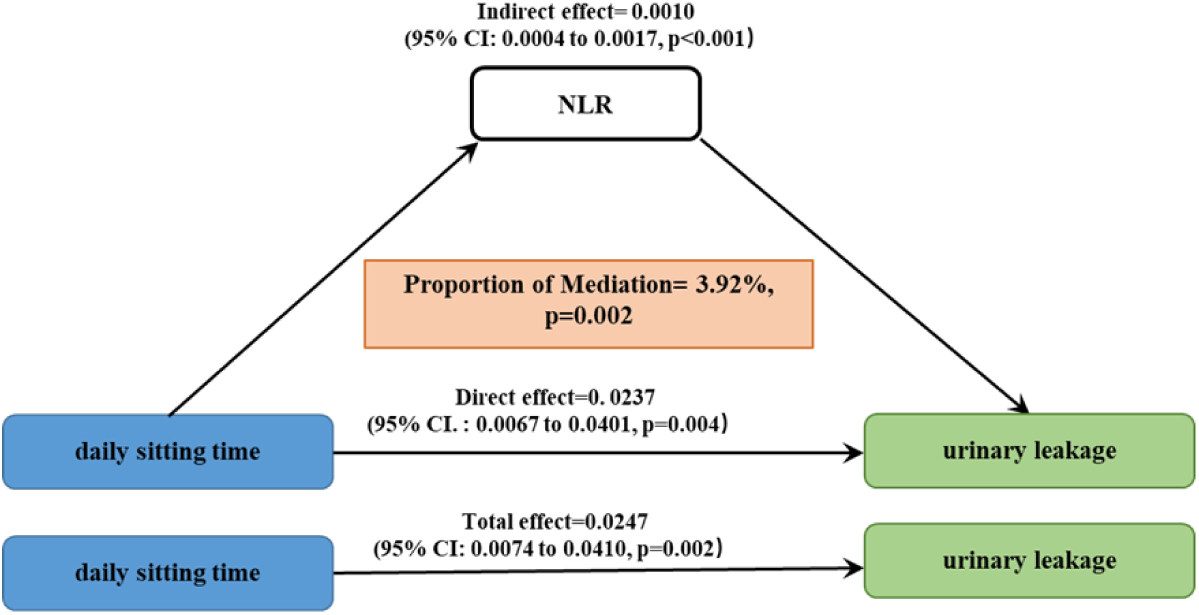
Path diagram of the mediation analysis of NLR on the relationship between DST and UL.

**Supplementary Fig.2.**
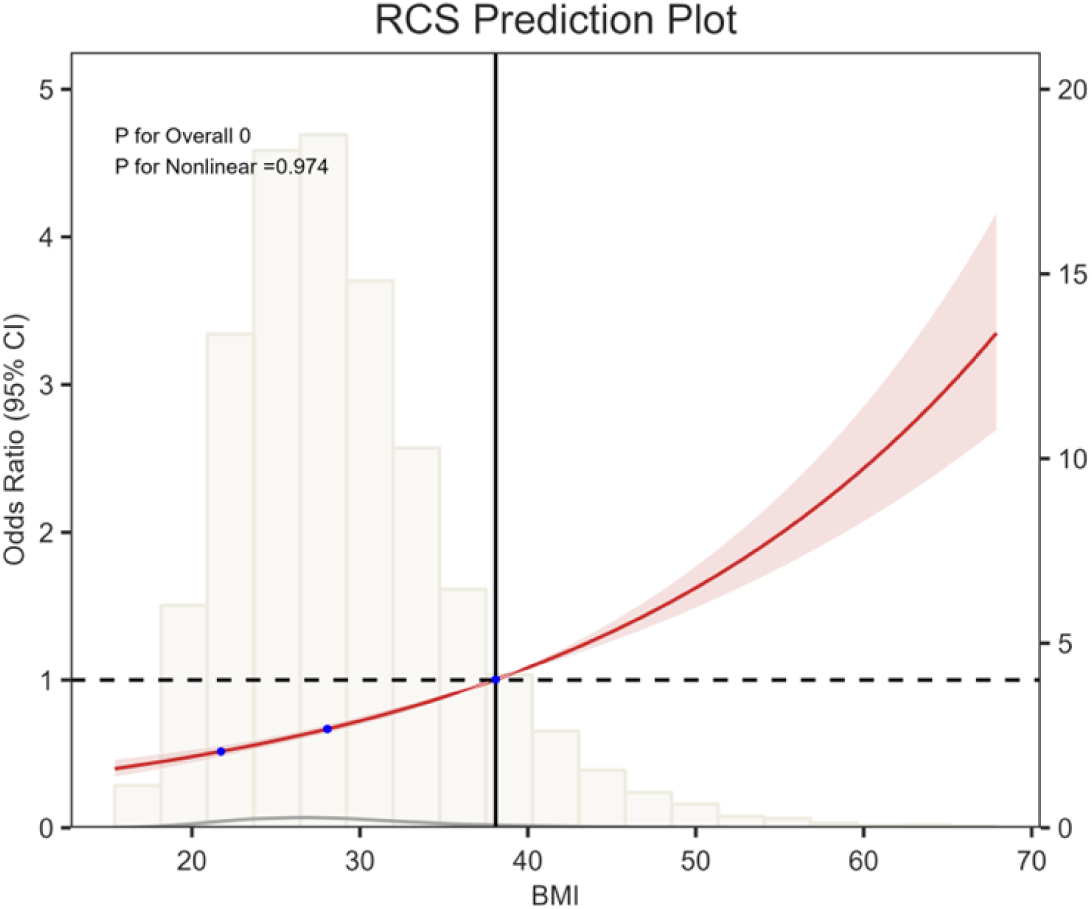
RCS revealed the association between BMI and the risk of UL caused by sedentary behavior.

